# Dihydropyridine Calcium Channel Blocker-induced Prescribing Cascades: Signal Detection using High-throughput Sequence Symmetry Analysis

**DOI:** 10.64898/2026.05.15.26353346

**Authors:** Priyanka J. Kulkarni, Asinamai Ndai, Shailina Keshwani, Kayla M. Smith, Jaeyoung Choi, Michael Luvera, Julia Hunter, Shannon Wright, Julia Hetzel, Carl J. Pepine, Stephan Schmidt, Earl J. Morris, Steven M. Smith

**Affiliations:** Department of Pharmaceutical Outcomes and Policy, College of Pharmacy, University of Florida, Gainesville, Florida, USA; Center for Integrative Cardiovascular and Metabolic Disease, University of Florida, Gainesville, Florida, USA; Center for Drug Evaluation and Safety (CoDES), Gainesville, Florida, USA; Division of Cardiovascular Medicine, University of Florida College of Medicine, Gainesville, Florida, USA; Center for Pharmacometrics and Systems Pharmacology, Department of Pharmaceutics, College of Pharmacy, University of Florida, Gainesville, Florida, USA

**Author notes:** **Correspondence:** Steven M. Smith, Address: 1889 Museum Road, Suite 6300, DSIT Building Gainesville, FL 32611.

**Keywords:** Dihydropyridine calcium channel blockers, prescribing cascades, high-throughput sequence symmetry analysis, Medicare

## Abstract

**Background:** Dihydropyridine calcium channel blockers (DHP-CCB) are widely prescribed antihypertensives whose adverse effects may trigger unnecessary prescribing of additional medications, termed prescribing cascades (PC). We aimed to identify potential DHP-CCB-induced PCs using high-throughput sequence symmetry analysis (HTSSA).

**Methods:** Using Medicare claims data (2011-2020), we identified new users aged ≥66 years with continuous enrollment ≥360 days before and ≥180 days after DHP-CCB initiation. We screened for initiation of 446 “marker” drug classes within ±90 days of DHP-CCB initiation. Sequence ratios compared marker drug initiation after versus before DHP-CCB initiation. Adjusted sequence ratios (aSR), accounting for prescribing trends over time, were calculated with 95% CIs >1 considered statistically significant. Clinical experts classified statistically significant signals as potential PCs through consensus.

**Results:** Among 388,862 DHP-CCB initiators (mean age 76.6 ± 7.5 years; 62.5% women, 92.3% with hypertension), 82 of 446 marker drug classes had significantly elevated aSRs, of which 24 were classified as potential PCs. Strongest signals ranked by highest aSR included other systemic hemostatics (aSR 2.99; 95% CI, 1.10-8.16), other nasal preparations (aSR 1.99; 95% CI, 1.47-2.70), and drugs used in erectile dysfunction (aSR 1.85; 95% CI, 1.27-2.70). Other clinically relevant signals, ranked by number needed to harm (lowest to highest), included sulfonamides (NNTH 104; 95% CI, 98-111), electrolyte solutions (NNTH 216; 95% CI, 196-241), and osmotically acting laxatives (NNTH 710; 95% CI, 540-1056).

**Conclusion:** Potential PCs identified in this Medicare cohort reflected known and underrecognized adverse effects of DHP-CCBs. Further studies are needed to evaluate the clinical consequences of these PCs.

## Introduction

Calcium channel blockers (CCBs) are widely used antihypertensive agents recommended by most hypertension guidelines as first-line therapy.^1–6^ They exert their effect by binding to L-type long-acting voltage-gated calcium channels, thereby inhibiting calcium influx into cardiac tissue, vascular smooth muscles and pancreatic cells, leading to a reduction in blood pressure.^7,8^ Both dihydropyridine and non-dihydropyridine CCBs are effective monotherapy, and are often used in combination with other antihypertensive agents.^4,8^

Dihydropyridine CCBs (DHP-CCBs) are preferred agents in hypertension as they reduce cardiovascular risk.^8–14^ They are generally well-tolerated, but can cause side effects such as headaches, flushing, hypotension, and peripheral edema.^4^ Incident lower-extremity edema, for example, has been frequently documented with DHP-CCBs, with risk increasing according to dose and duration of therapy.^15^ In some instances, providers may misinterpret this edema as a new medical condition and subsequently prescribe a loop diuretic to manage this adverse event (AE), triggering a phenomenon known as a PC.^9,15–23^ Prior studies have identified real-world evidence of this PC, reporting more than two-fold greater loop diuretic initiation after, compared with before, initiation of a high-dose DHP-CCB.^9^

PCs are more common in older adults compared to younger adults due to a higher comorbidity burden and polypharmacy.^16^ Although the DHP-CCB-loop diuretic PC is well-known, evidence on other unknown PCs are limited in the United States (US).^24–28^ Sequence symmetry analysis is a pharmacovigilance tool being used in studies involving administrative healthcare databases to identify potential drug-induced PCs.^29^ Identification of unknown DHP-CCB-induced PCs could aid in informing safe prescribing practices, reducing healthcare costs and improving patient quality of life by avoiding potentially inappropriate use of medications.

## Methods

### Data Source

We used Medicare administrative claims data from 2011 to 2020. Medicare is a federal health insurance program designed to provide coverage for Americans aged ≥65 years and those <65 years with specific disabilities or end-stage renal disease.^30^ Specifically, we used a 5% sample of 2011-2015 Medicare claims and a 15% sample of 2016-2020 Medicare claims. The samples were enriched with 1 million Florida Medicare beneficiaries (2011–2015) and 100% Florida beneficiaries (2016–2020). We restricted our analysis to beneficiaries enrolled in fee-for-service plans including inpatient hospital (Part A), outpatient (Part B), and prescription drug coverage (Part D) to allow more complete capture of patient-level pharmacy and medical claims for healthcare services received by beneficiaries. The study was considered exempt by the University of Florida institutional review board and we used the STROBE (Strengthening the Reporting of Observational Studies in Epidemiology) guideline to ensure appropriate reporting.

### Design

We used HTSSA to identify potential DHP-CCB-induced PCs. HTSSA is a hypothesis-free pharmacovigilance approach that employs a case-only study design to assess the temporality of an “index” drug initiation (herein, DHP-CCB) relative to “marker” drugs (i.e., all other medication classes), hereafter referred to as DHP-CCB-marker class dyads. We included new users of DHP-CCB who had their first DHP-CCB fill between 2011-2019, were aged ≥66 years at DHP-CCB initiation and had continuous insurance coverage ≥360 days before and ≥180 days after DHP-CCB initiation (Supplemental Figure S1). We excluded patients who initiated other antihypertensive drug classes on the same day as DHP-CCB initiation.

For the high-throughput screening, we hierarchically grouped marker drugs into medication classes using Anatomical Therapeutic Classification (ATC) codes. The ATC classification system, maintained by the World Health Organization, classifies medications into groups at 5 levels, where level 1 indicates the broad anatomical group (n=14), level 4 indicates the chemical subgroup/drug class, and level 5 indicates the specific drug/chemical substance (n≈5000) (Supplemental Table S1). We identified the first claim for any marker drug within a given ATC Level 4 class. If an individual filled multiple unique medications within a level 4 category during the study period, we only considered the date of the first medication filled within the ATC level 4 category. We restricted the marker drug initiation window to within ±90 days of DHP-CCB initiation for the primary analysis, to capture cascades likely driven by acute AE onset. The window was extended to ±180 days in sensitivity analyses to evaluate the robustness of results and allow for the identification of AEs and subsequent potential PCs cascades with longer induction periods. For each DHP-CCB-marker dyad, we excluded patients who initiated a DHP-CCB and given marker drug on the same day.

All DHP-CCB-marker class dyads were evaluated using PSSA. Analyses were completed iteratively until all ATC level 4 categories were evaluated. Baseline characteristics (age, sex, calendar year of DHP-CCB initiation, Charlson Comorbidity Index, specific DHP-CCB medication, and other comorbidities) of DHP-CCB initiators were measured in a 360-day lookback period and used for stratified analysis.

### Statistical Analysis

For each unique DHP-CCB-marker class dyad, we calculated the crude sequence ratio (cSR) as the number of patients who initiated marker drug after DHP-CCB initiation divided by the number of patients who initiated marker drug before DHP-CCB initiation. An aSR ≈ 1 indicates no causal relationship between the prescribing patterns; conversely, excess initiation of a marker class after the index drug relative to before index drug initiation would result in an aSR >1 and be consistent with a potential PC. To account for secular prescribing trends (e.g., increasing or decreasing use of DHP-CCB or marker drug over time), we derived the null-effect sequence ratio for each DHP-CCB-marker class dyad. Briefly, the null-effect ratio is the expected sequence ratio in the absence of any causal relationship between marker and index drug, based on population level prescribing trends.^31^ We then estimated an adjusted sequence ratio (aSR) with a 95% confidence interval (CI) by dividing the cSR by the null effect ratio for each dyad to adjust for background prescribing trends. All aSRs with a lower CI limit >1 were considered statistically significant under the assumption that no within-person time-varying effect exists.

The unique DHP-CCB-marker class dyad was represented graphically by plotting the distribution of the timing of marker class initiation (in 10-day intervals) relative to DHP-CCB initiation for each exposure window (±90 days and ±180 days). Additionally, for each statistically significant dyad, we estimated excess risk among the exposed and the corresponding number needed to harm (NNtH) within a year. Excess risk among the exposed was calculated as the difference between the number of patients who initiated the marker class after DHP-CCB initiation and those who initiated the same marker class before DHP-CCB initiation, divided by total number of DHP-CCB initiators, standardized to a rate per 1000 person-years. NNtH was calculated as the inverse of the excess risk among the exposed, i.e., the number of DHP-CCB initiators needed for one additional patient to experience a PC within that marker class.

### Classification of signals

We systematically conducted a pharmacological review of the statistically significant signals to differentiate potential PCs from false positive signals. False positives could result from detection bias (i.e., a new condition identified with a corresponding new medication initiated during routine monitoring of DHP-CCB), disease progression (i.e., new medication initiated to treat worsening of the underlying cardiovascular disease), therapeutic escalation (i.e., escalation of therapeutic regimen from 1^st^ line to 2^nd^ line agents and so on which is unrelated to the DHP-CCB indication), or reverse causation (i.e., reduced DHP-CCB initiation following marker class initiation such as late-stage chemotherapy). These signals were considered as not indicative of potential PCs and classified as “Other” in the review process.

The pharmacological review was conducted in 2 stages. First, pharmacy trainees were trained by study investigators in the use of secondary data sources namely PubMed/Medline, drug monographs, package inserts, and drug information data sources such as Micromedex and SIDER to assess the underlying mechanisms of signals to support classification as potential PCs or not.^32^ Two pharmacy trainees reviewed each significant signal and assigned it an initial classification, based on supporting literature and the process described above. Two pharmacists, with clinical expertise in medication and patient safety, independently classified each signal using material developed by pharmacy trainees and ad-hoc literature evaluations when needed. In the event of disagreement between the reviewers, consensus was sought through discussion with the study team that included a physician and a clinical pharmacologist.

All analyses were conducted in SAS statistical software version 9.4 (SAS Institute, Cary, NC), visualized in RStudio version 2025.05.1+513 and Tableau (2023.06.2 + 561).

## Results

We identified 388,862 patients who initiated a DHP-CCB during the study period (Table 1). Of these, 62.5% were women. The mean ± SD age of the patients was 76.6 ± 7.5 years, with 45.4% aged 65-74 years and 37.4% aged 75-84 years. Most patients were non-Hispanic Whites (79.4%) and had a Charlson Comorbidity Score of ≥5 (76.7%). Amlodipine was the most frequently prescribed DHP-CCB (95.5%).

**Table 1:**
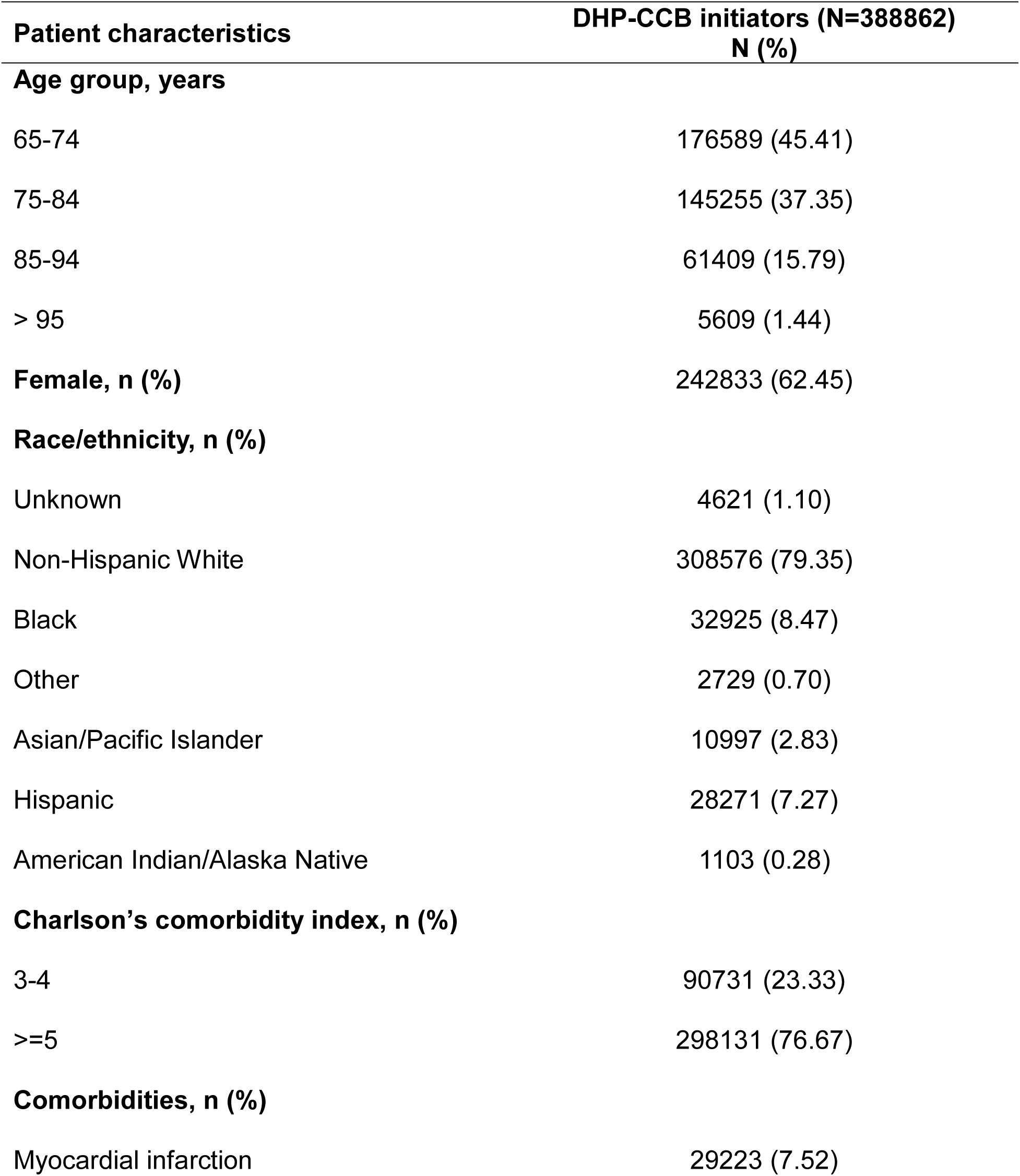
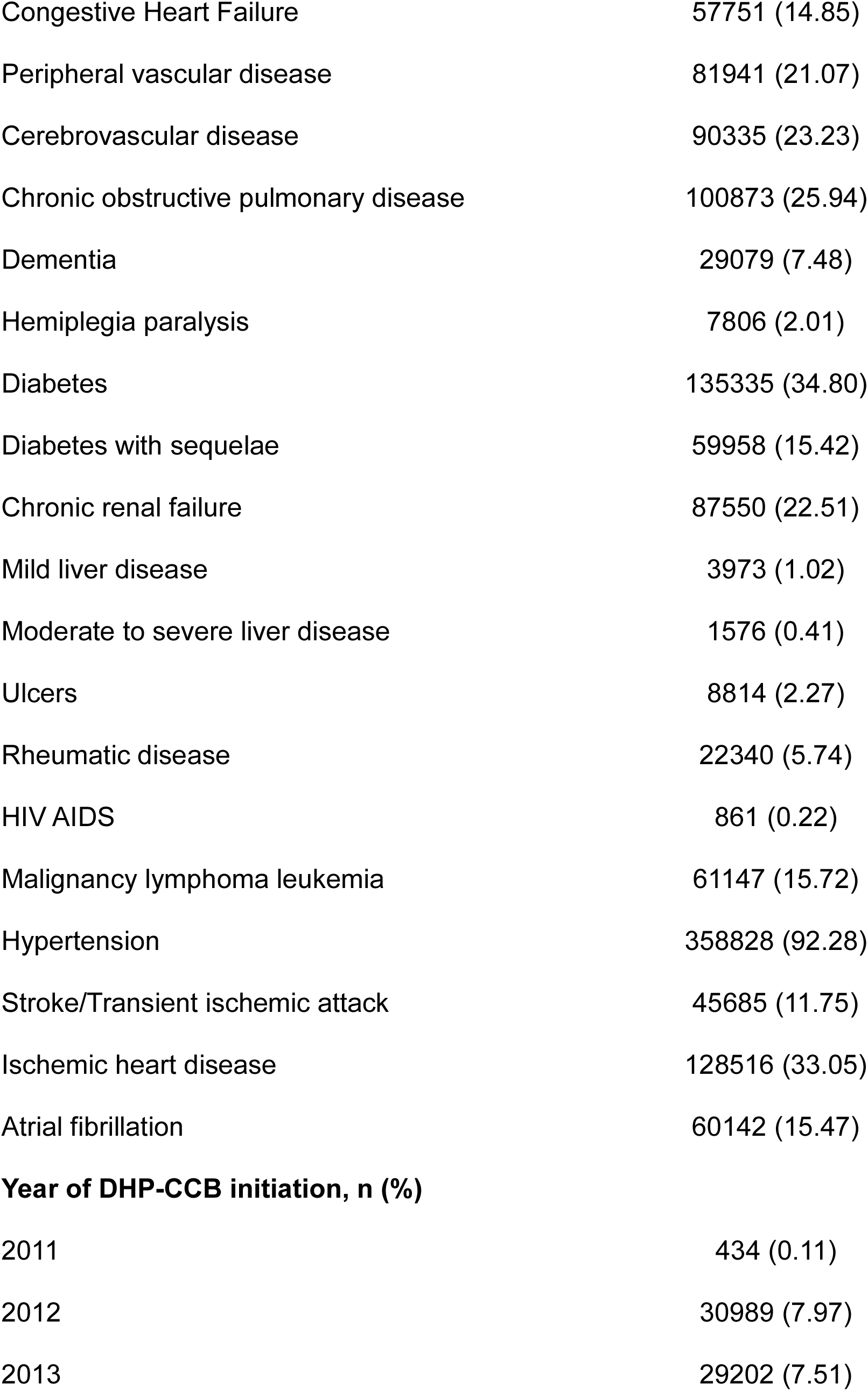
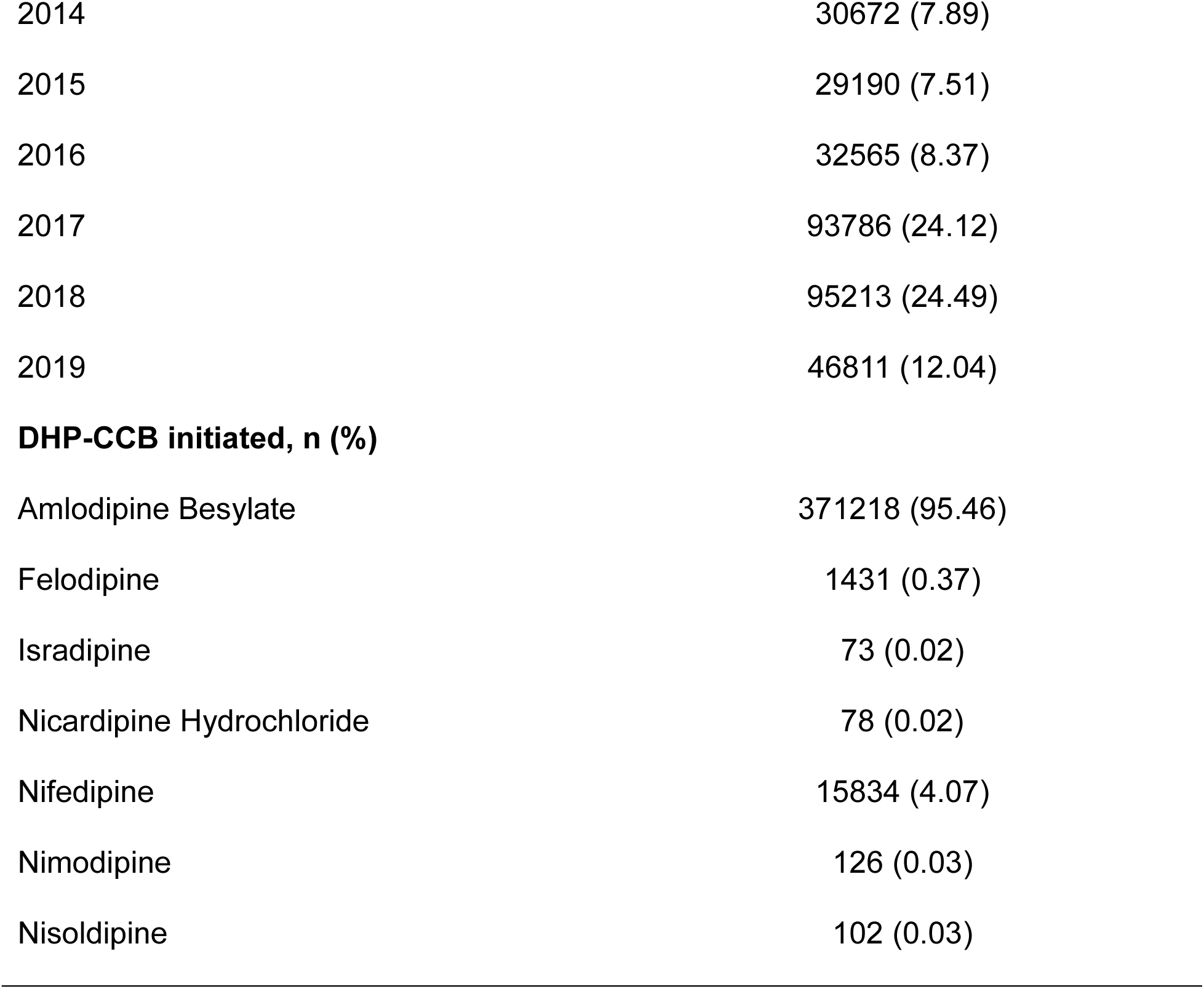
Baseline characteristics of DHP-CCB initiators included in the cohort.

Of 446 marker classes assessed, 82 statistically significant signals (aSR 95% CI wholly >1) were identified in the 90-day primary analysis (complete list is shown in Supplemental Table S2). After clinical review, 24 (29.3%) signals were classified as potential (biologically plausible) PCs. Of the 82 statistically significant signals identified in the ±90-day analysis, 51 (62%) were also significant in the ±180-day sensitivity analysis (Supplemental Table S3). All high-throughput screening results are interactively displayed at https://public.tableau.com/app/profile/cvmedlab/vizzes.

PCs classified as clinically plausible are displayed in Figure 1, ranked by decreasing aSR. Classification of these signals at the ATC 1 level found that 29% belonged to the nervous system, 21% belonged to the alimentary tract and metabolism, 17% to respiratory system, 13% to blood and blood forming organs, 8% to dermatologicals, 4% to systemic hormonal preparations, 4% to genito-urinary system and sex hormones and 4% to cardiovascular system.

**Figure 1:**
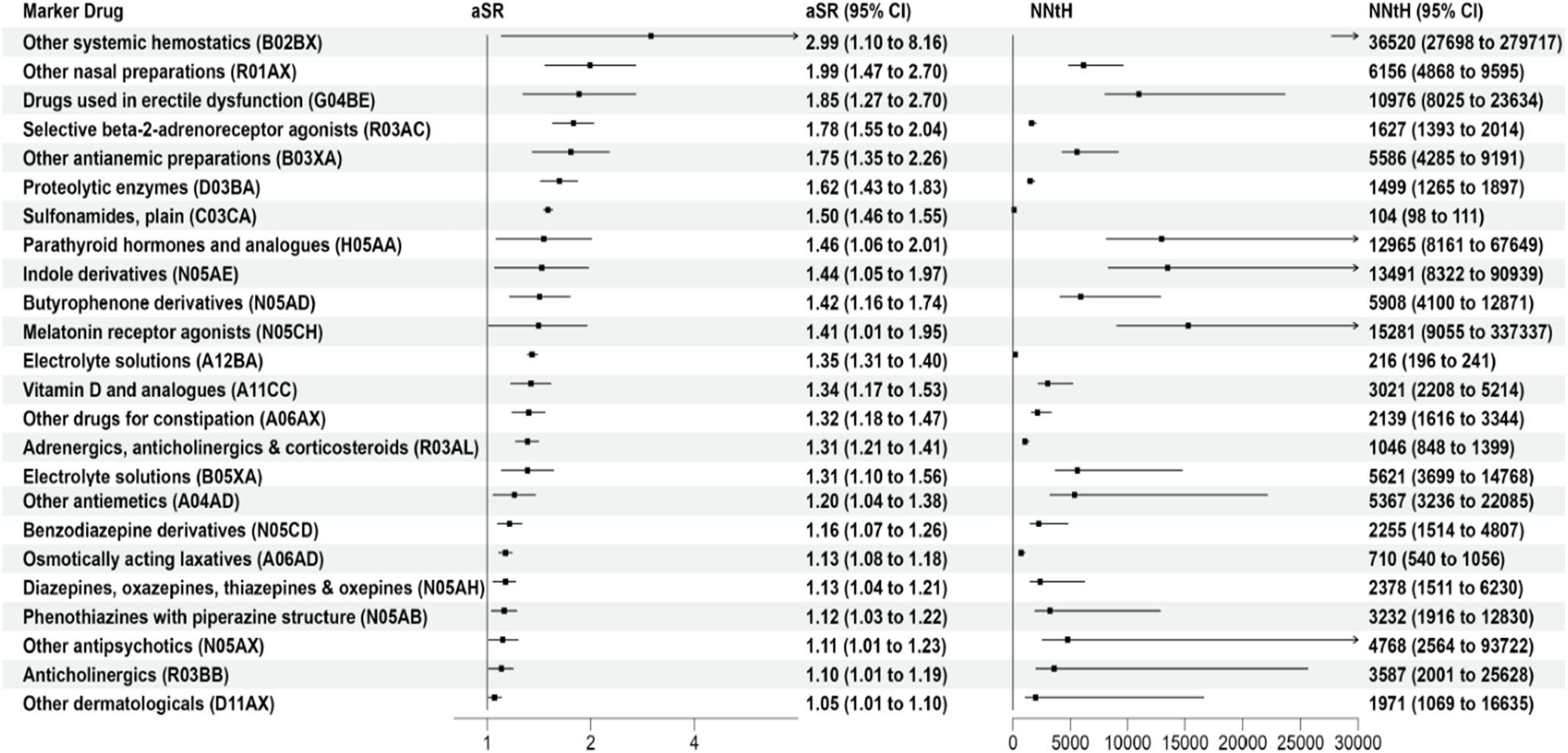
Clinically plausible potential prescribing cascades ranked by adjusted sequence ratio (aSR) NNtH: Number needed to harm; aSR, CI

Signals were ranked using (1) aSR, a measure of the magnitude of the signal, and (2) NNtH, a measure of the excess risk of marker drug prescription in those exposed to DHP-CCBs. The top 3 potential PCs ranked by aSR were: other systemic hemostatics (aSR 2.99, 95% CI. [1.10,8.16]) including the drugs romiplostim, eltrombopag and fostamatinib; other nasal preparations (aSR 1.99, 95% CI [1.47, 2.70]) which includes ipratropium bromide; and, drugs used in erectile dysfunction (aSR 1.85, 95% CI [1.27, 2.70]) which includes sildenafil and vardenafil. Ranking by the NNtH (lowest to highest), the top 3 potential PCs were: sulfonamides (NNtH 104, 95% CI [98, 111]) which includes furosemide, bumetanide and torsemide; electrolyte solutions (NNtH 216, 95% CI% [196, 241]), which includes potassium salts; and, osmotically acting laxatives (NNtH 710, 95% CI [540, 1056]) which includes magnesium oxide, lactulose, sodium sulfate, macrogol (i.e., polyethylene glycol) and sodium phosphate.

We conducted stratified analyses among patients with and without chronic kidney disease (CKD) and diabetes mellitus (DM), as these indications are associated with the use of antianemic drugs. The signal was found to be significant irrespective of CKD or DM status (Supplemental Table S4), indicating plausibility of the DHP-CCB-antianemic agents PC.

Figure 2 summarizes the statistically significant DHP-CCB-marker class dyads by NNtH and aSR and highlights those that were classified as potential PCs.

**Figure 2:**
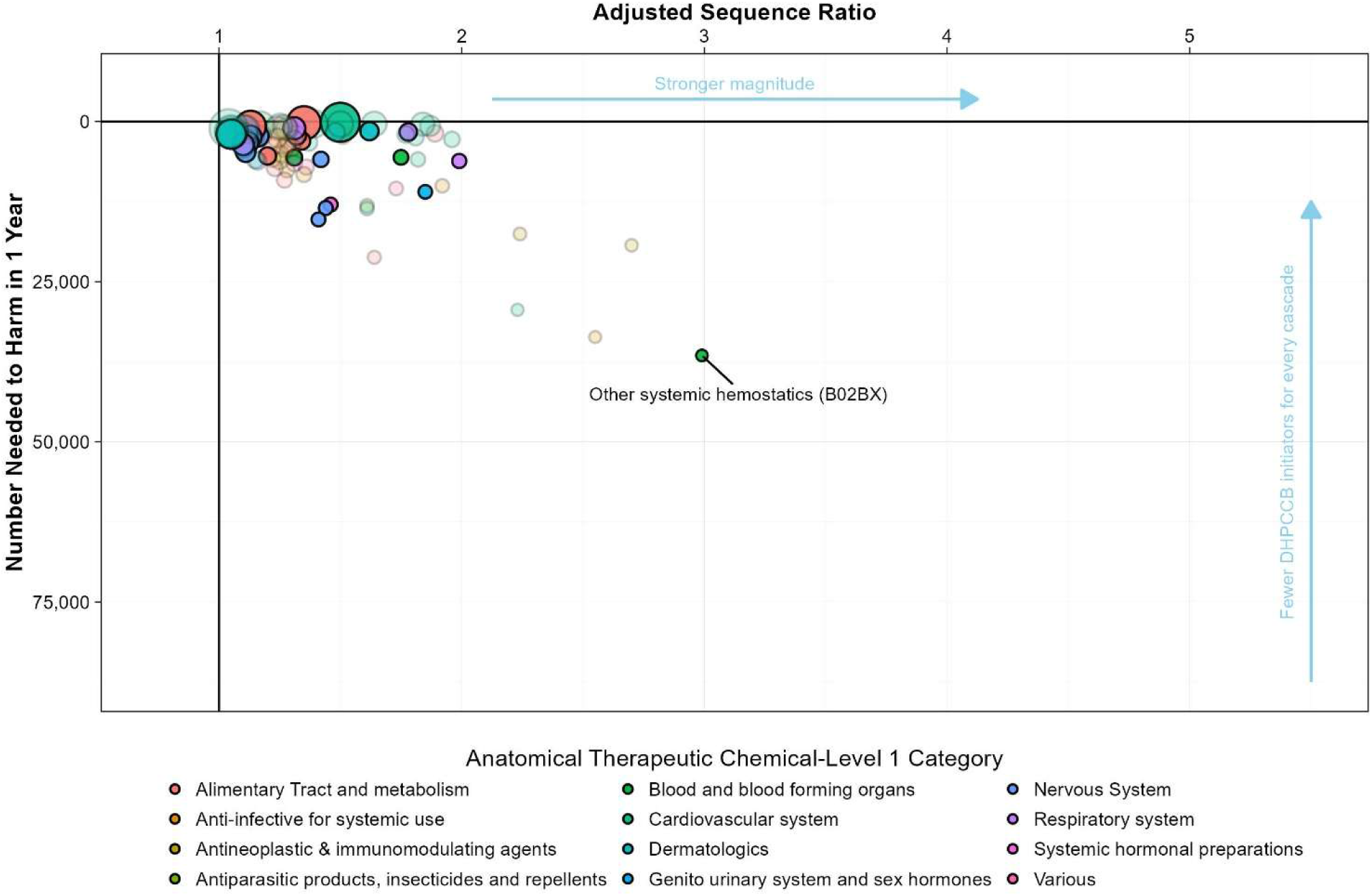
Significant DHP-CCB-marker class dyad signals by adjusted sequence ratio and naturalistic number needed to harm. Dyads are grouped (color-coded) at the Anatomical Therapeutic Chemical Level 1 category. The faded dots are dyads classified as “other” while the colored dots are dyads classified as potential prescribing cascades weighted by the total number of prescriptions. All results from the high-throughput screening are displayed interactively at https://public.tableau.com/app/profile/cvmedlab/vizzes

## Discussion

Using population-based HTSSA of Medicare data (2011-2020), we identified 82 signals suggestive of real-world DHP-CCB-induced PCs, of which 24 were deemed clinically plausible after review. Some of the observed signals, for example, the DHP-CCB-loop diuretic PC mediated by DHP-CCB-induced peripheral edema, have been well described ^9,16,25^ and, in our study, was associated with the lowest NNtH (104 95% CI [98,111]), reflecting in part its high incidence. However, we also observed signals that may indicate previously unknown or underrecognized PC signals.

Systemic hemostatics- romiplostim, eltrombopag and fostamatinib- had the strongest association (aSR, 2.99; 95% CI, 1.10-8.16). Although the underlying mechanism remains uncertain, drug-dependent antiplatelet antibodies have been documented with amlodipine causing thrombocytopenia.^33–35^ Evidence also suggests that DHP-CCBs may inhibit platelet activation through vasodilation-mediated prostacyclin release.^36^ These pathways may explain the elevated use of systemic hemostatics in our cohort. Interestingly, fostamatinib is known to cause systemic elevations in blood pressure, highlighting a potentially significant cascade loop that could be prevented by timely discontinuation or substitution of the DHP-CCB.^37^

Excess use of anti-anemic drugs- erythropoietin and darbepoetin alfa after DHP-CCB initiation could be explained by reduced hemoglobin, RBC or hematocrit levels. DHP-CCB use is associated with hemoglobin loss, potentially due to impaired platelet aggregation and decreased vasoconstriction.^38–46^. While this effect is prominent in patients with DM and CKD, these diseases are also independently associated with anemia.^47–49^ Our stratified analyses, however, confirm that this PC is significant irrespective of CKD or DM status.

Excess prescribing of ipratropium bromide after DHP-CCB initiation may suggest treatment for nasal congestion related to the vasodilatory effects of DHP-CCBs.^50–52^ Case reports have also linked DHP-CCBs to acute respiratory distress syndrome (ARDS) with proposed mechanisms including arterial vasodilation, reflex tachycardia, endothelin-1-mediated reduction in surfactant secretion causing alveolar collapse, and precapillary vasodilation leading to interstitial edema.^53–57^ These mechanisms may explain the observed signals for selective beta-2 adrenoceptor agonists (i.e., salbutamol, formoterol, indacaterol, olodaterol), anticholinergics like ipratropium bromide, tiotropium bromide, and combination inhalers.

Hypertension increases the risk of erectile dysfunction, particularly among treated individuals, although the effect of antihypertensive therapy itself versus disease severity remains uncertain.^58^ Evidence for a class effect of antihypertensives on erectile dysfunction is inconsistent.^59–63^ However, a Dutch community pharmacy cohort study reported a significant association between DHP-CCBs and erectile dysfunction (aSR of 1.76 [1.68, 1.84])^25^ which is consistent with our findings for sildenafil and vardenafil (aSR [95% CI]: 1.85 [1.27, 2.7]).

Case reports also suggest that amlodipine may induce gingival hyperplasia.^64,65^ Proposed mechanisms include TGF β1 upregulation, impaired collagenase activity, reduced folic acid uptake, aldosterone inhibition, and increased keratinocyte growth factor regulation.^65^ These mechanisms may explain the excess prescribing of proteolytic enzymes such as collagenase after DHP-CCB initiation. DHP-CCBs have additionally been associated with cutaneous reactions like pruritus, urticaria and alopecia^32,66^ potentially explaining the observed prescribing of dermatological agents such as topical minoxidil.

DHP-CCBs may also affect bone mineral density and parathyroid hormone (PTH) regulation. Proposed mechanisms include calciuria, loop diuretic-induced volume depletion, and direct stimulation of parathyroid tissue.^67–69^ This could explain the increased use of PTH analogs such as teriparatide (aSR [95%CI] 1.36 [1.07, 1.72]). DHP-CCB-induced hypotension may further increase fall and fracture risk, prompting osteoporosis treatment, although detection bias is also possible. Notably, the teriparatide signal remained significant in the 180-day sensitivity analysis, supporting its potential clinical plausibility. Altered calcium homeostasis may likewise explain the increased prescribing of Vitamin D analogs such as calcitriol.^70^

We observed increased use of antipsychotics including prochlorperazine, haloperidol, ziprasidone, olanzapine, and risperidone, consistent with observational evidence linking antihypertensive use to increased risk of mood disorder-related hospitalization.^71^ DHP-CCBs, particularly amlodipine have also been associated with insomnia potentially explaining PCs involving lorazepam, and ramelteon.^72^

Gastrointestinal adverse effects DHP-CCBs, including constipation, nausea and diarrhea, are well recognized, although their mechanisms are poorly understood. Animal studies have demonstrated dose-dependent inhibition of intestinal motility with amlodipine and nifedipine,^73,74^ while a non-interventional study reported increased constipation risk with amlodipine use.^75^ These findings suggest reduced gastrointestinal peristalsis and delayed gastric emptying.^75^ Experimental studies have also implicated L-type CCBs in gastroesophageal reflux disease and aldosterone synthesis.^76–78^ Because aldosterone regulates potassium homeostasis, its inhibition can contribute to electrolyte distrurbances.^78^ A randomized double-blinded trial additionally demonstrated increased natriuresis with nifedipine use.^79^ Together these mechanisms may explain the observed signals for laxatives (e.g. magnesium oxide), constipation therapies (e.g. lubiprostone), antiemetics (e.g. dronabinol), and electrolyte solutions (e.g. sodium salts).

This study has several strengths. We used a nationally-representative sample of older adults with hypertension from Medicare fee-for-service data, improving generalizability within elderly populations. The large sample size also enhanced statistical power. In addition, the case-only design inherently controlled for time-invariant confounders (e.g. sex, age, race/ethnicity) and certain within-person biases, including disease severity, while the analyses adjusted for prescribing trends over time. However, several limitations should be considered. The primary analysis used a ±90-day exposure window, extended to 180 days in sensitivity analyses which may have missed PCs with longer latency periods. Future studies should consider longer assessment windows to detect slower-developing signals. Signal classification also relied on clinical expertise and literature-informed review, introducing potential misclassification bias despite consensus-based adjudication. Furthermore, findings may not generalize to younger or uninsured populations. Lastly, all signals identified in this study are hypothesis-generating and require further validation in robust retrospective cohort studies.

In summary, we identified both known and potentially novel PCs induced by DHP-CCBs using HTSSA in a population of Medicare beneficiaries. While the findings are hypothesis-generating, they provide useful information to clinicians on optimizing anti-hypertensive therapy while minimizing risks from potentially inappropriate medication use in older adults with hypertension.

## Data Availability

The study was conducted using CMS Medicare Fee-For-Service claims databases, pursuant to a data use agreement between University of Florida and CMS that prevents the sharing of data entrusted to the University of Florida. However, qualified researchers can obtain such data directly from CMS. The SAS code used in this study is available from the Corresponding Author upon reasonable request.

https://public.tableau.com/app/profile/cvmedlab/vizzes

## Acknowledgements

None

## Author Contributions

Priyanka J. Kulkarni contributed to the study design and drafted the manuscript. Steven M. Smith, Earl J. Morris, and Scott M. Vouri conceived and designed the study and secured funding. Earl J. Morris, Priyanka J. Kulkarni, Asinamai M. Ndai, Shailina Keshwani, Scott M. Vouri, Kayla Smith, and Steven M. Smith were responsible for the statistical analysis and visualization of the data. All authors made substantial contributions to the interpretation of the data and results, reviewed, provided critical revisions, and approved the final manuscript.

## Supplemental Material

Tables S1–S4

Figure S1- S2

